# Prescribed footwear and orthoses are not prophylactic in preventing lower extremity injuries in military tactical athletes. A systematic review with meta-analysis

**DOI:** 10.1101/2021.06.07.21258508

**Authors:** Scott L. Paradise, Joshua R. Beer, Chris A. Cruz, Ken M. Fechner, Andrew J. MacGregor, John J. Fraser

**Affiliations:** Primary Care Sports Medicine Fellowship, Naval Hospital Camp Pendleton, Oceanside, CA, USA; United States Navy Medicine Readiness and Training Command Guam, Agana, GU, USA; Uniformed Services University of the Health Sciences, Bethesda, MD, USA; United States Navy Medicine Readiness and Training Unit, Marine Corps Recruit Depot Parris Island, Parris Island, SC, USA; United States Navy Medicine Readiness and Training Command Camp Pendleton, Oceanside, CA, USA; Directorate for Operational Readiness & Health, Naval Health Research Center, 140 Sylvester Road, San Diego, CA, USA

## Abstract

**Introduction:** Military members are exposed to high cumulative physical loads that frequently lead to injury. Prescribed footwear and orthoses have been used to prevent injury. The purpose of this systematic review with meta-analysis was to assess if prescribed prophylactic footwear or foot orthoses reduced lower extremity injury risk in military tactical athletes.

**Methods:** MEDLINE, Embase, Web of Science, CINAHL, SportDiscus, and DTIC databases were searched for randomized controlled trials published at any time that compared foot orthoses or prescribed footwear (to include shock-absorbing insoles and socks) to a placebo intervention or a no-treatment control. Methodological quality was assessed and numbers of injuries, population at risk, and the duration of the study epoch were extracted and relative risk (RR) calculated. An *omnibus* meta-analysis was performed assessing all prescribed footwear and orthoses intervention studies, with subgroup analyses conducted on studies with similar interventions [i.e., basketball athletic shoes; athletic shoes (prescribed by foot type); foot orthoses; shock-absorbing insoles; socks; tropical combat boots].

**Results:** Of 1,673 studies identified, 22 studies were included. Three of eight studies that employed orthoses demonstrated significantly reduced overuse injuries compared to no treatment controls (RR range: 0.34-0.68); one study showed neoprene insoles significantly decreased overuse injuries (RR: 0.75). There were no other significant effects in the individual studies, and no protective effects observed in the *omnibus* meta-analysis or in the component sub analyses.

**Conclusions:** Prescribed footwear and orthoses do not appear to have a prophylactic effect on lower quarter MSKI in military members and cannot be recommended at this time.

## INTRODUCTION

Musculoskeletal injuries (MSKI) are common during military training and operations and can adversely affect medical readiness, warfighter performance, and mission accomplishment. MSKI, primarily of the lower quarter, were leading reasons for outpatient medical encounters in 2019 in the Military Health System.[1,2] These injuries substantially contribute to medical-related attrition and the multibillion per-annum direct and indirect healthcare cost for active military members and veterans.[2] The etiology of many non-battle related MSKI are repetitive, microtraumatic overuse injuries resulting from high intensity exercises and cumulative loads incurred primarily during marching and running.[3,4] The substantial burden imposed by MSKI warrants in-depth assessment of preventive interventions used to mitigate these injuries.

Foot orthoses, shock-absorbing insoles, and other prescribed footwear have been used for the prevention of overuse injuries in athletes and military recruits.[5–7] These interventions alter lower quarter biomechanics by attenuating ground reaction forces, distributing plantar pressures, and altering kinematics during functional tasks.[5,8–10] However, there is mounting scrutiny regarding the effectiveness of footwear prescription for running-related prophylaxis.[11] While prescribed orthoses or footwear may mediate potential intrinsic risk factors, such as foot phenotype, this does not necessarily translate to reduction of injury. With the types and volume of exposure, unique hazards, and a “mission first” culture unique to the military that precludes care-seeking,[12] it is unclear whether prophylactic orthoses or prescribed footwear would be effective in MSKI prophylaxis in this unique population.

The physical demands placed on military tactical athletes during training are inherently different than those incurred by their civilian counterparts. Military members are exposed to high cumulative physical loads resulting from frequent and high intensity training, often with little respite. Given the unique exposures specific to the military, it is unclear whether foot orthoses, shock-absorbing insoles, or prescribed footwear would be protective against MSKI. While prior systematic reviews have evaluated whether prophylactic footwear or ankle-foot orthosis prescription were able to reduce injury in the civilian population,[6,7,14] none at the time of writing have evaluated MSKI prophylaxis in the military specifically. Therefore, the purpose of this systematic review with meta-analysis was to assess if prescribed prophylactic footwear or foot orthoses reduced lower extremity injury risk in military tactical athletes.

## METHODS

The protocol for this study was registered *a priori* in PROSPERO (CRD42020183403, http://bit.ly/CRD42020183403). The Preferred Reporting Items for Systematic Reviews and Meta-Analyses (PRISMA)[15] and A MeaSurement Tool to Assess systematic Reviews version 2 (AMSTAR 2)[16] were used to guide study reporting.

### Eligibility Criteria

Studies were eligible for inclusion if they were randomized controlled trials that compared foot orthoses or prescribed footwear (to include shock-absorbing insoles and socks) to a placebo intervention or a no-treatment control. All studies must have reported the inclusion of military tactical athletes, the burden (number, rate, or proportion) of lower extremity injuries for both the intervention and control groups, the at-risk population size, and the duration of the study epoch. If the required information could not be ascertained from the published study, the corresponding authors were contacted. Studies were excluded if they were systematic reviews or retrospective studies, if the interventions were not randomized, or if the data was not available for extraction.

### Search Strategy

A research librarian was consulted to develop the search strategy. The search strategy, comprised of MeSH terms, is detailed in the Supplemental Table 1 (https://doi.org/XXXXXXX). The searches were limited to records in English, the native language of the study team, published at any time of inquiry. MEDLINE, Embase, Web of Science, CINAHL, SportDiscus, and the Defense Technical Information Center (DTIC) databases were queried on January 10, 2020. DTIC (https://discover.dtic.mil/) serves as the research repository of the U.S. Department of Defense. Records were organized and duplicates were removed using Rayyan QCRI, an application used to facilitate study selection for systematic reviews (https://rayyan.qcri.org/). Two reviewers (SLP and JB) independently reviewed each record by title and then abstract for inclusion. A third author (KF) resolved any disagreements. Study selection is detailed in the PRISMA flowsheet (Figure 1).

**Table 1.** Risk of bias assessment for included studies

|  | Andrish 1974 | Bonanno 2018 | Bensel 1976 | Bensel 1983 | Esternan 2005 | Finestone 1999 | Franklyn-Miller 2011 | Gardner 1988 | Hesarikia 2014 | Knapik 2009 | Knapik 2010a | Knapik 2010b | Larsen 2002 | Mattila 2011 | Milgrom 1985 | Milgrom 1992 | Mundermann 2001 | Schwellnus 1990 | Sherman 1996 | Smith 1985 | Van Tiggelen 2009 | Withnall 2006 |
| --- | --- | --- | --- | --- | --- | --- | --- | --- | --- | --- | --- | --- | --- | --- | --- | --- | --- | --- | --- | --- | --- | --- |
| Random sequence generation | ? | + | - | ? | + | ? | + | - | + | - | - | + | + | + | ? | ? | ? | ? | - | ? | ? | + |
| Allocation concealment | ? | + | ? | ? | ? | + | + | - | + | - | - | + | + | + | ? | ? | ? | ? | - | ? | ? | + |
| Blinding of participants and personnel | - | + | - | - | - | - | - | + | - | ? | ? | ? | ? | ? | ? | - | ? | - | - | ? | ? | - |
| Blinding of outcome assessment | ? | + | ? | ? | ? | - | ? | + | ? | + | + | + | + | + | ? | ? | ? | ? | ? | ? | ? | ? |
| Incomplete outcome data | - | + | ? | ? | + | - | + | + | ? | ? | + | + | + | + | - | + | - | + | - | - | ? | + |
| Selective outcome reporting | + | + | + | + | ? | + | + | ? | - | + | + | + | + | + | + | + | - | + | + | ? | + | ? |
| Other | + | + | + | + | + | + | + | + | + | + | + | + | + | + | + | + | - | + | - | + | + | + |

**Figure 1.**
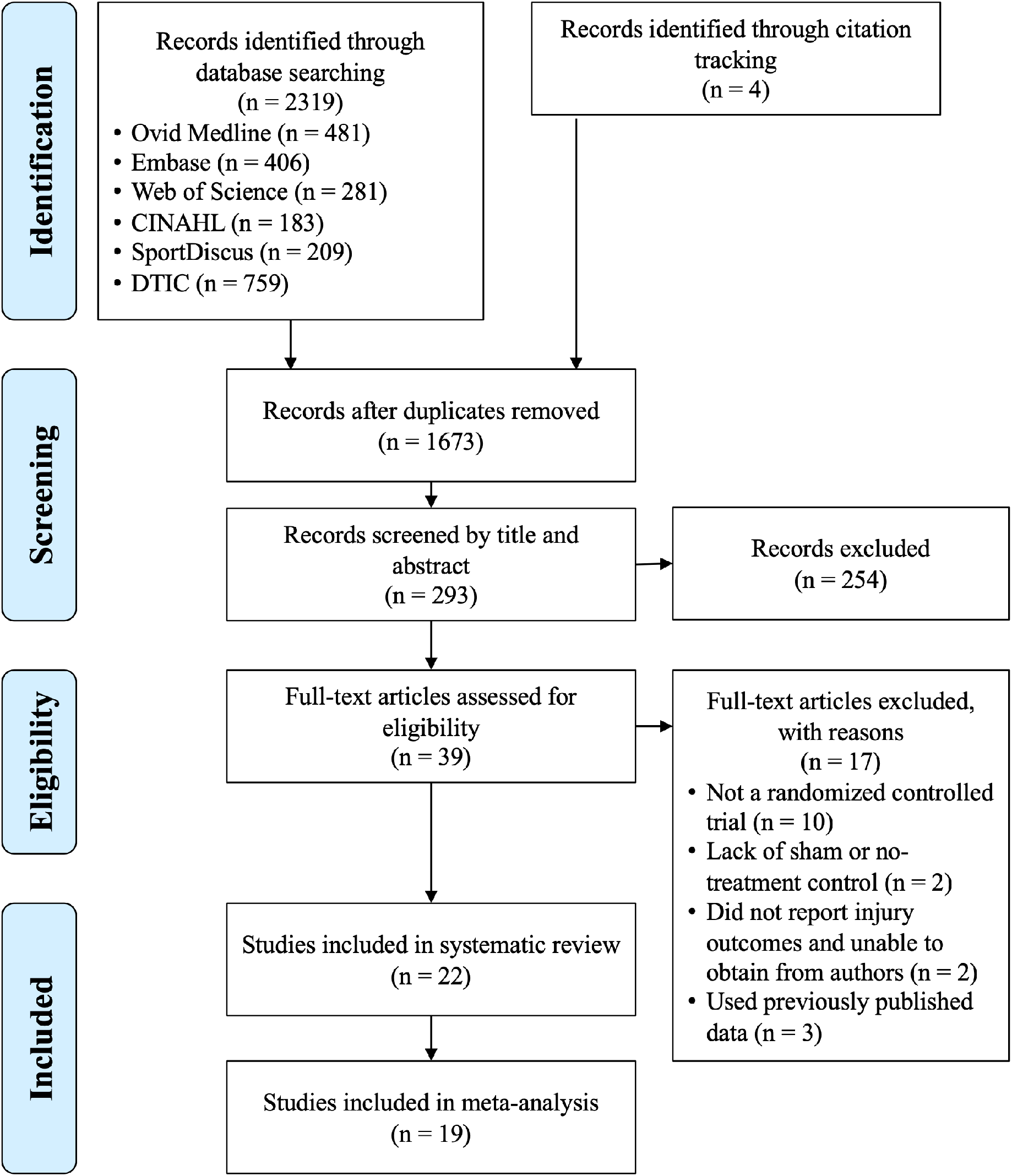
PRISMA flow diagram of study selection process. CINAHL, Cumulative Index of Nursing and Allied Health Literature; DTIC, Defense Technical Information Center.

### Data Extraction

Two reviewers (SLP and JRB) independently assessed each report for extractable data using the Cochrane Collaboration Data Collection Form for RCTs and non-RCTs. The number of injuries, the number of the population at risk, and the duration of the study epoch were extracted. Studies that reported incidence or prevalence measures were reverse calculated to extract count data.

Study characteristics pertaining to participant demographics, trial setting, method of randomization, and intervention characteristics are reported in Supplemental Table 2 (https://doi.org/XXXXXXX). Any disagreements were resolved by consensus. If consensus could not be achieved, a third author (KF) resolved any disagreements. Cumulative incidence of lower quarter MSKI for both intervention and control groups were calculated using the number of injuries during the study epoch and the population size at the time of allocation. Calculations of relative risk with 95% confidence intervals (CI), attributable risk (AR), and number needed to treat (NNT) were used to assess prophylactic effects for each study.

### Risk of Bias Assessment

Risk of bias for each study were assessed using the Cochrane Collaboration tool.[17] Each study was assessed in seven domains: random sequence generation, allocation concealment, blinding of participants and personnel, blinding of outcome assessment, incomplete outcome data, selective outcome reporting, and other. Each domain was independently rated by two authors (SLP and JRB) as high risk of bias, low risk of bias, or marked as unclear. Reviewers resolved disagreements by consensus, and a third author (KF) was consulted to resolve disagreements if needed. A study was judged to have overall high risk of bias if at least one domain was rated as having high risk, or if there were concerns in multiple domains that substantially lowered confidence in its results.[17]

### Synthesis Methods

An *omnibus* meta-analysis was performed assessing all prescribed footwear and orthoses intervention studies, with subgroup analyses conducted on studies with similar interventions [athletic shoes (basketball); athletic shoes (prescribed by foot type); foot orthoses; shock-absorbing insoles; socks; tropical combat boots]. Pooled outcomes were calculated using the Mantel-Haenszel method with the Hartung-Knapp adjustment for random effects models.[18,19] Heterogeneity was assessed using the *I*^*2*^ and χ^2^ statistics and conclusions were contextualized according to risk of bias. *I*^*2*^ statistics were interpreted as suggested by Higgins and colleagues, with higher values indicating greater heterogeneity.[20] A leave-one-out sensitivity analysis and Baujat plot[21] were used to diagnose specific studies contributing to heterogeneity. Reporting bias was assessed with a funnel plot and Egger’s statistic to evaluate symmetry. Data synthesis was performed using the ‘meta’ package (version 4.18-0) for R version 4.0.3 (The R Foundation for Statistical Computing, Vienna, Austria).

## RESULTS

### Study Selection

The search strategy yielded 1673 records after duplicates were removed (Figure 1). Four additional records were identified through cross-referencing citations.[22–25] Of these, 39 full-text records were assessed for eligibility, and 22 included in the systematic review. Three studies were excluded as these were not randomized intervention trials.[26–28] Simkin et al.[29] and Finestone et al.[30] met inclusion criteria, however they were extensions of the studies conducted by Milgrom et al. in 1985[31] and 1992[30], respectively, and were excluded. Sherman et al. [32] was included in the systematic review but excluded from the meta-analysis because the study epoch was not specified. Mundermann et al.[22] was included in the systematic review but excluded from the meta-analysis due to insufficient data.

### Study Characteristics

Supplemental Table 2 (https://doi.org/XXXXXXX) details the extracted study characteristics that include the setting, population at risk, intervention, comparison, time at risk, and injury outcome of interest. Eight studies evaluated the effect of biomechanical orthoses on overuse injury incidence in military recruits.[10,31,33–38] Six studies evaluated the effect of shock-absorbing inserts.[9,22,25,32,39,40] One study evaluated the effect of shock-absorbing heel cups with and without heel-cord stretches.[24] Six studies evaluated the prophylactic effect of prescribed footwear,[30,41–45] and one study evaluated the effect of padded and double-layered socks.[23]

Three trials employed cluster randomization to enhance participant blinding.[9,23,32] The remaining trials randomized each participant. It was otherwise impossible for trials to maintain blinding of orthosis or insert among military units who often live and train together in close confines, so four studies made use of a sham insert to minimize bias.[9,10,34,40] Studies included predominantly male participants (range: 56% to 100%), a finding attributed to conscription practices[37,38] or training site demographics.[44] Trials typically occurred over the course of an initial recruit training course or initial period of service which ranged from 6 weeks[45] to 6 months.[38] Andrish et al.[24] specified only that training occurred over the summer. Since “Plebe Summer” at the US Naval Academy is 7 weeks in duration, it was assumed that this was the study duration. Sherman et al.[32] also did not specify the epoch length, but did mention that it occurred during Army basic training that was 8 weeks long at the time of the study.

Five studies described a protocol to confirm stress fractures or other injuries with the use of radiographs[9,38] and Technetium bone scans.[30,31,34] One study confirmed stress fractures with magnetic resonance imaging.[35] Bonanno et al.[10] was the only study that referenced a previously published research protocol and specified use of standardized clinical assessments. Three studies counted any injury severe enough to cause a limitation in training.[35,38,40] One study required a limitation in training of at least one day,[39] and one study only counted injuries severe enough to limit training for a period of three days.[33] The remaining studies either specified a clinical exam by a healthcare provider or study team member, review of diagnostic codes from a patient record, or did not specify diagnostic criteria.[22–25,32,36,37,41,41,43–45]

### Risk of Bias Assessment

Only one study was rated as having low risk of bias (Table 1).[10] The remaining studies were unclear or had high risk of bias. Four studies were rated as low risk in the majority of the assessment categories.[10,37,38,44] Lack of blinding of participants, or the failure to report blinding, was the primary threat to validity across the majority of studies. To a lesser extent, uncertainty pertaining to blinding of outcome data due to lack of granularity in methodological reporting was also common.[22–25,30–33,35,36,39–42] Finally, there were a considerable number of trials that were rated as unclear or high risk of bias pertaining to allocation concealment and random sequence generation.[9,22–25,30–34,39,41–43,45] Milgrom et al.[31] observed substantial dropout in the orthosis intervention group (21.0%) due to discomfort and analyzed only the remaining participants, which posed a substantial source of attrition bias. Esterman et al.[33] observed substantially low compliance in injured recruits introducing differential bias, and was excluded from the meta-analysis.

### Study Findings

#### Orthoses

Three of the eight studies that employed orthoses demonstrated significantly reduced overuse injuries when compared to no treatment controls (RR range: 0.34 to 0.68).[31,35,37] Finestone et al.[34] reported a significant protective effect by combining semi-rigid polypropylene and soft polyurethane intervention arms. When analyzed individually, both polypropylene (RR: 0.59) and polyethylene (RR: 0.61) orthoses had wide 95% CIs that were statistically non-significant. Lastly, the intervention group in the study conducted by Hesarikia et al.[36] was approximately twice as likely to experience an injury while wearing orthoses compared to the no-treatment controls (AR: 14.2 per 1000 person-weeks, NNT Harm: 9). There were no further significant findings in studies assessing prophylactic orthoses.

#### Shock-Absorbing Insoles

Two studies reported a reduction in overuse injuries in recruits provided with shock-absorbing insoles compared to no treatment controls.[25,39] Of these, one study showed neoprene insoles significantly decreased overuse injuries (RR: 0.75, NNT Benefit: 15), but not stress fractures.[39] While Smith[25] reported improved injury rates for US Coast Guard recruits who were provided either Spenco (9.5%) or Poron (8.7%) insoles compared to no treatment controls (29.2%), calculations of RR with 95% CI using extracted data were non-significant [Spenco: 0.29 (0.06, 1.26); Poron: 0.29 (0.06, 1.26)]. There were no further significant findings in studies assessing prophylactic insoles.

#### Prescribed Footwear

Studies of prescribed athletic footwear by arch height in military recruits reported no significant effects.[43–45] However, our calculations of RR using extracted data suggest that the prescribed footwear may have actually have had a significant, but small, increase in injury risk in Air Force recruits (RR: 1.11, NNT Harm: 29).[45] In a study assessing prescribed tropical combat boots compared to standard issue leather boot controls in Marine recruits, the intervention group were reported to have significantly higher occurrence of retrocalcaneal bursitis, but not other overuse injuries.[41] Calculations of RR based on overall injury occurrence suggest no significant effect. In a follow-on study conducted with Army recruits,[42] the intervention group that wore the tropical combat boot had significantly more injuries (RR: 1.39, NNT Harm: 17) than the standard leather boot control group.[42] In a study assessing the prophylactic effects of padded polyester socks or a two-sock system (a thin, inner polyester sock worn under a thick, outer cotton-wool sock) compared to the standard issue uniform sock, padded polyester socks prevented blisters (RR: 0.53, NNT Benefit: 3), an outcome the authors used as a surrogate for knee joint overuse injury.[23]

### Results of Syntheses

There were no significant protective effects observed in the *omnibus* meta-analysis or in the component analyses assessing pooled effects of athletic shoes prescribed by foot type, foot orthoses, shock-absorbing insoles, socks, or tropical combat boots compared to controls. There was considerable heterogeneity observed in the *omnibus* synthesis (Figure 2). In the subanalyses of similar interventions, heterogeneity ranged from low (shock absorbing insoles) to substantial and considerable (foot orthoses, tropical boots). Subanalyses of interventions with the highest degree of heterogeneity also had the fewest number of studies included (socks, tropical boots), with exception of foot orthoses. While there were studies that contributed a substantial degree of heterogeneity identified on the Baujat plot (Figure 3), the sensitivity analysis found that omission of these studies would only minimally reduce total heterogeneity (Supplemental Table 3, https://doi.org/XXXXXXXX).

**Figure 2.**
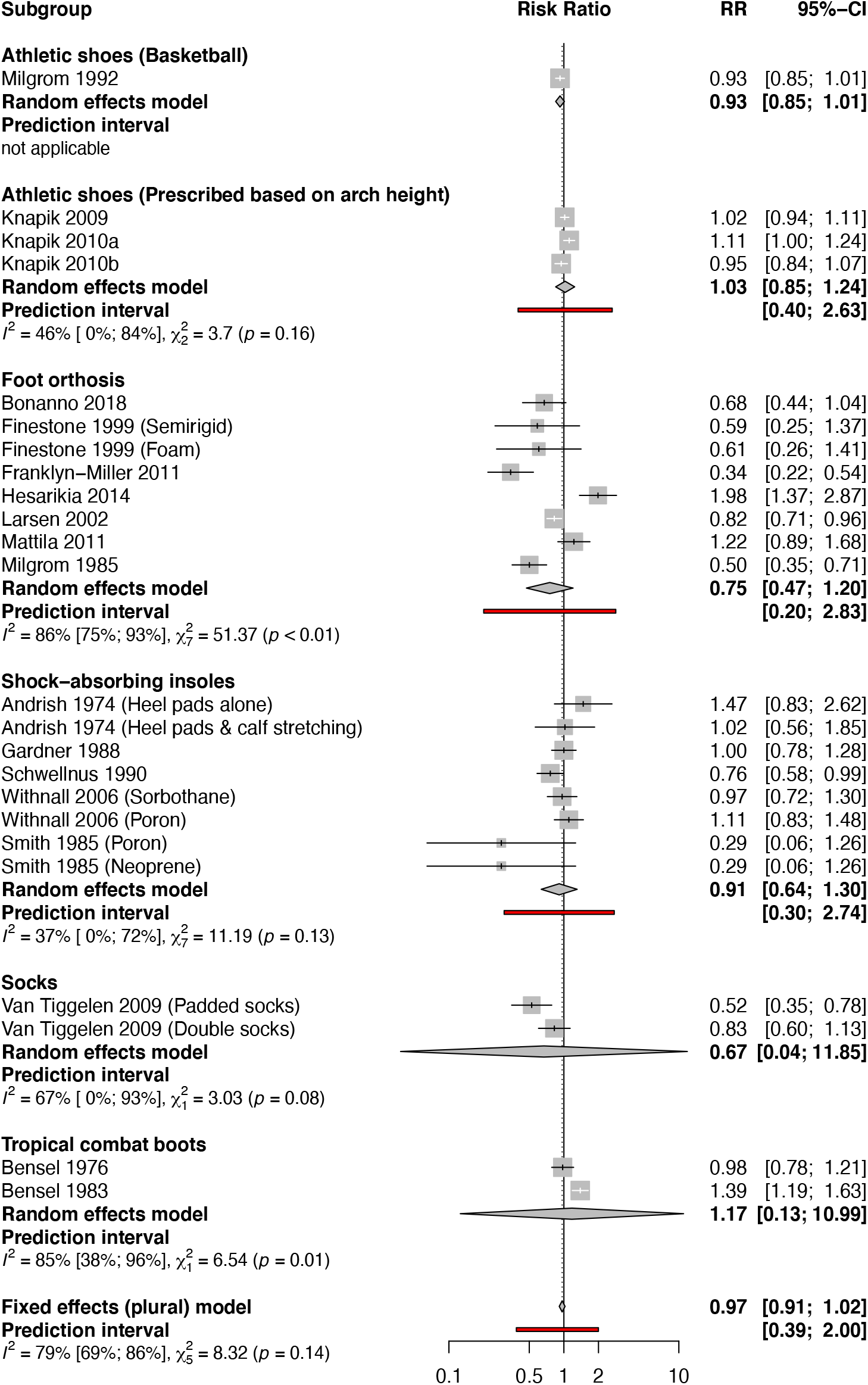
Meta-analyses of lower extremity injuries by intervention

**Figure 3.**
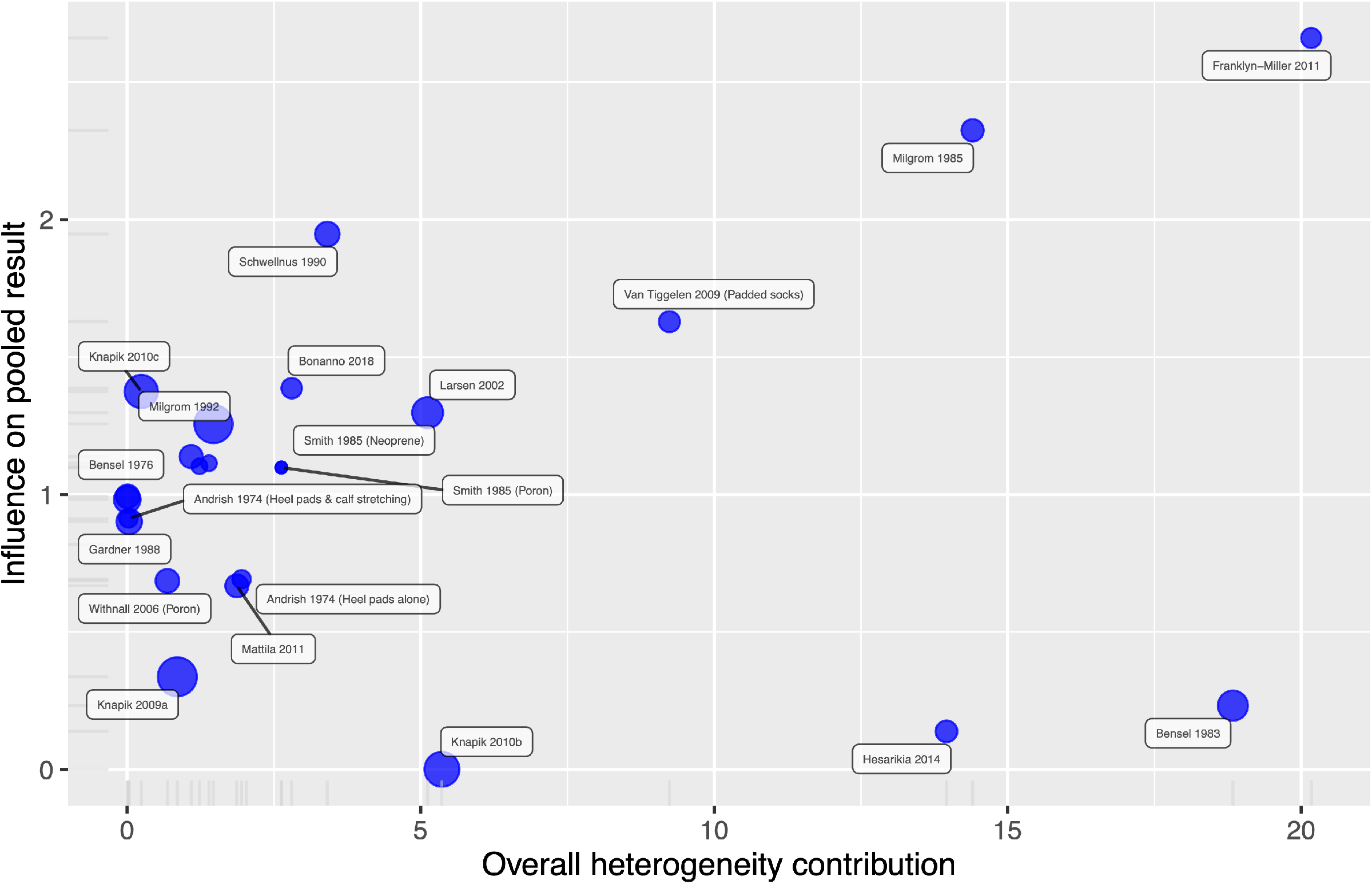
Baujat plot of study heterogeneity.

### Reporting Bias

Significant asymmetry was found in the Eggers test, which is visualized in the funnel plot (Figure 4). At the apex of the funnel, there was greater symmetry in the studies with the lowest standard errors. As standard error increased, it appears that there was bias toward studies that demonstrated protective effects.

**Figure 4.**
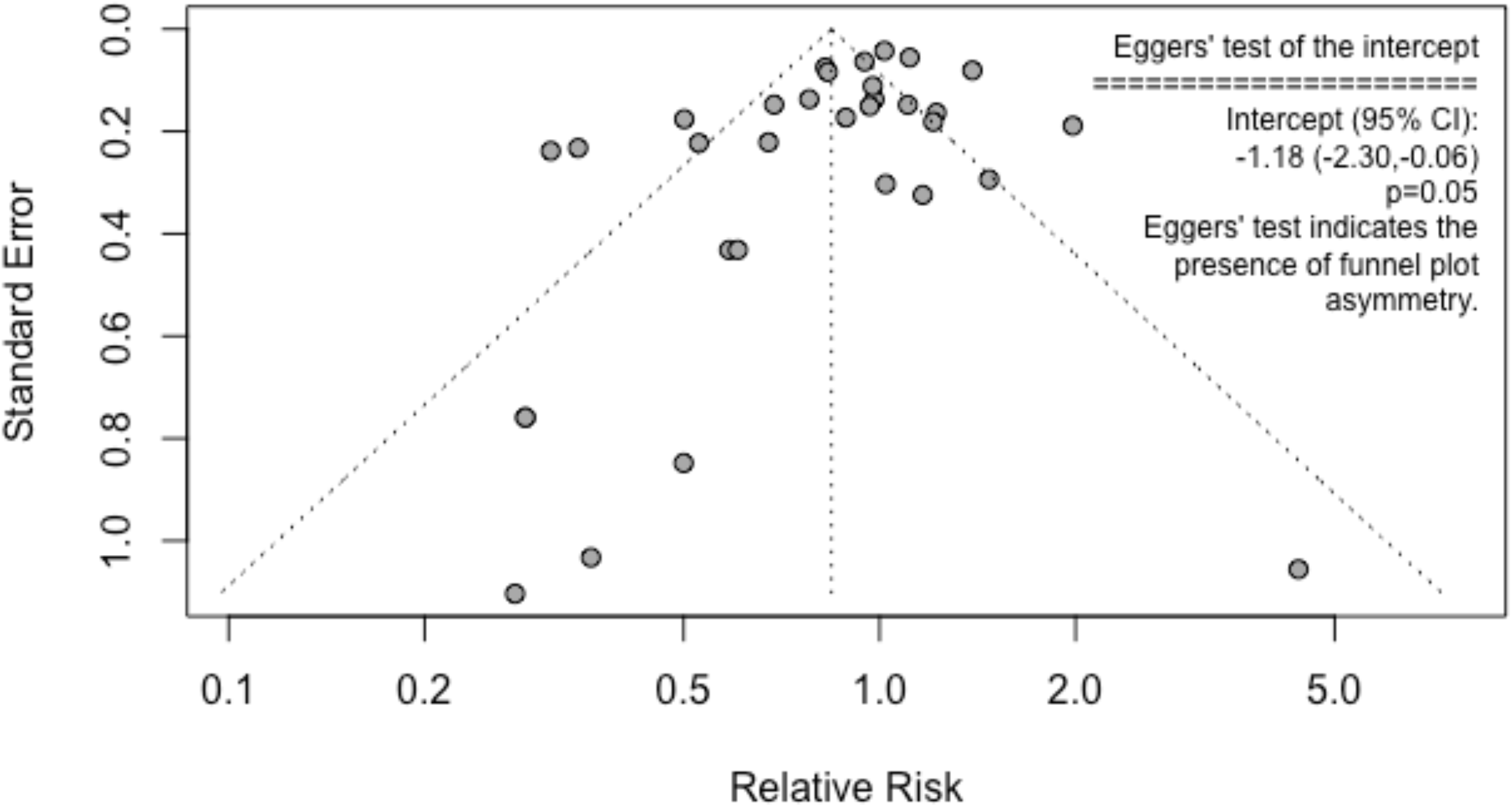
Funnel plot assessing reporting bias.

## DISCUSSION

The primary finding of this study was that orthosis, insole, or footwear prescription did not have a pooled prophylactic effect on lower quarter MSKI in military members. Since most of the included studies that prescribed interventions did not consider the individual characteristics or needs of the military member, the widespread application of the non-specific interventions employed in these studies cannot be recommended at this time. Due to the substantial heterogeneity and the risks of bias observed across the reviewed studies, these findings should be interpreted based on the limitations of these trials.

Our findings agree with those reported by Knapik et al.[46], Yeung et al.[7], and Rome et al.[6] that found inconclusive evidence for the use of prescribed orthoses, insoles, or footwear for the prevention of injury. Among these studies, only the synthesis conducted by Knapik et al.[46] employed a military-only study population in the aggregation of their previous three studies of Army, Marine Corps, and Air Force recruits.[43–45] While our findings pertaining to prescribed shock-absorbing insoles agree with the those found in the meta-analysis conducted by Bonanno el al.,[14] our findings pertaining to the lack of prophylactic effects of foot orthoses diverge.[14] Their analysis found that orthoses were effective in preventing overall MSKI. [14] In the meta-analysis of injury type, this was found to be limited to stress fractures and not soft tissue injuries.[14] The divergence in our findings may have been a result of combining all injuries from each study prior to inclusion in the analyses.

From a clinical perspective, widespread and non-specific prescription of orthoses, insoles, or footwear cannot be recommended at this time. Bullock et al.[3] similarly concluded that there was insufficient evidence to recommend prescribed footwear based on arch height, the use of shock-absorbing insoles, or replacement of footwear at regular intervals for the prevention of injury. This should not be interpreted to preclude the utilization of these interventions for specific clinical indications identified during examination of trained medical professionals. From a research perspective, our findings raise more questions than answers. There is a need for high quality prevention studies using contemporary research methods, specifically those outlined in the CONSORT guidelines.[47] Furthermore, it is unclear if policies such as the obligatory use of Berry Amendment compliant shoes, which are regulated to be domestically manufactured and issued based on foot type in US military recruits,[48,49] has an effect on injury and warrants future investigation.

There are limitations to this study. We utilized cumulative incidence measures at the time of allocation for calculations of relative risk. While this measure is consistent with the intention to treat principle, it does not account for attrition due to administrative reasons, which may have biased the results. While it can be assumed there was equity in both groups leading to non-differential bias, this is an assumption. We looked at overall injury burden and did not investigate if these interventions were protective against specific types of injuries. It is plausible that specific findings may have become non-significant by employing this approach. Lastly, we used non-peer reviewed research reports to mitigate the effects of publication bias. While this may have affected methodological quality of these studies, these studies[41,42] were not dissimilar from other studies that were vetted by peer reviewers.

## CONCLUSIONS

Prescribed footwear and orthoses do not appear to have a prophylactic effect on lower quarter MSKI in military members. Since most of the included studies that prescribed interventions did not consider the individual characteristics or needs of the military member, the widespread application of the non-specific interventions employed in these studies cannot be recommended at this time. These findings should be tempered based on the limitations of the studies in this area.

### Support

No external financial support was received for this study. We greatly appreciate the assistance of Simona Konecna for her assistance in developing the search strategy and executing the search.

### Key Messages

- Musculoskeletal injury is common in military populations and leads to impaired medical readiness and large financial costs.
- Prescribed footwear and orthoses have been proposed as measures for lower limb injury prevention.
- In military populations, prophylactic footwear and orthoses do not appear to have a preventive effect on lower limb injury rates.
- Future preventive studies should utilize high-quality, contemporary methodologies.

## Supporting information

Supplemental Table 1. Search strategy by database.

Supplemental Table 2. Study characteristics.

Supplemental Table 3. Sensitivity analysis results.

## Data Availability

The datasets used and/or analysed during the current study are available from the corresponding author on reasonable request.

