## Supplemental Table 1. Search strategy by database. for "Prescribed footwear and orthoses are not prophylactic in preventing lower extremity injuries in military tactical athletes. A systematic review with meta-analysis"

| Ovid MEDLINE | 1 aerospace medicine/ or military medicine/ or naval medicine/ or submarine medicine/ (49187)  2 armed conflicts/ or afghan campaign 2001-/ or gulf war/ or iraq war, 2003-2011/ or korean war/ or september 11 terrorist attacks/ or vietnam conflict/ or warfare/ or war exposure/ or war-related injuries/ (29207)  3 military personnel/ (38824)  4 hospitals, military/ (5188)  5 exp Military Facilities/ (202)  6 Military Nursing/ (2247)  7 ((military adj recruit$) or (nav$ adj recruit$) or (army adj recruit$) or (marine adj recruit$) or (infantry adj recruit$)).ti,ab. (1849)  8 (military or navy or army or (air adj force) or (special adj force$) or marines or ship or ships or naval or (service adj members) or (service adj personnel) or (armed adj forces) or submarine$ or corpsman or corpsmen or (coast adj guard) or aviator$ or (surface adj warfare) or (air adj crew) or (special adj operation$) or soldier$ or airman or airmen or (flight adj crew) or (military adj recruit$) or (naval adj recruit$) or (navy adj recruit$) or (army adj recruit$) or (corps adj recruit$) or (military adj forces) or (military adj force) or (marine adj corps) or (active adj duty) or cadet$ or (military adj deployment)).ti,ab. (83064)  9 1 or 2 or 3 or 4 or 5 or 6 or 7 or 8 (140809)  Annotation: All applicable military MeSH terms and keywords in title and/or abstract  10 foot orthoses/ (894)  11 shoes/ (6102)  12 exp orthotic devices/ (12739)  13 ((shoe$ adj insert$) or shoewear or (shoe adj wear) or pedorthic or orthose$ or ortose$ or orthotic or ortotic or footwear or (foot adj wear) or (shoe adj ortho$) or boots or (military adj boot$) or (infantry adj boots)).ti,ab. (8524)  14 10 or 11 or 12 or 13 (22103)  Annotation: All applicable orthotic MeSH terms and keywords in title and/or abstract.  15 9 and 14 (325)  Annotation: Military results set and Orthotic results set combined  16 tendinopathy/ or tendon entrapment/ or exp tenosynovitis/ (8525)  17 exp Athletic Injuries/ (26591)  18 exp ankle injuries/ (9591)  19 exp foot injuries/ (4264)  20 exp soft tissue injuries/ (5367)  21 exp tendon injuries/ (23689)  22 exp leg injuries/ (94191)  23 exp preventive medicine/ (35080)  24 back injuries/ or exp hip injuries/ (31391)  25 fasciitis/ or fasciitis, plantar/ (3299)  26 "sprains and strains"/ (5149)  27 cumulative trauma disorders/ (4280)  28 exp Musculoskeletal System/in [Injuries] (91671)  29 16 or 17 or 18 or 19 or 20 or 21 or 22 or 23 or 24 or 25 or 26 or 27 or 28 (260350)  Annotation: All applicable injury MeSH terms  30 ((injur$ adj athletic$1) or (injur$ adj overuse) or (injur$ adj soft adj tissue$1) or (injur$ adj tendon$1)).ti,ab. (633)  31 (injur$ adj (leg$1 or hip$1 or knee$1 or ankle$1 or foot or lower limb$1)).ti,ab. (1768)  32 (plantar fasciitis or (pain adj (heel or knee)) or shin splint$1 or tibial stress syndrome).ti,ab. (1725)  33 (preventive adj medicine).ti,ab. (5603)  34 30 or 31 or 32 or 33 (9717)  Annotation: All applicable injury keywords in title and/or abstract  35 29 or 34 (265504)  Annotation: Injury MeSH terms or Injury keywords  36 15 and 35 (133)  Annotation: All three sets combined (Military + Orthotic + Injuries)  37 limit 36 to english language (131)  Annotation: All three sets limited to English  38 15 not 37 (194)  Annotation: Articles from set #15 (Military and Orthotic) that were not included in set #35  39 limit 38 to english language (178) |
| --- | --- |
| DTIC R&E Gateway | ("foot ortho*" OR  "shoe insert*" or shoe-insert* or  "shoe insole*" or shoewear or shoe NEXT/1 wear OR orthotic* or ortotic*or orthose* or orthosis* OR footwear or  feet or  foot NEXT/1 wear OR "shoe ortho*" or boot or boots or "military boots" or "leg brace*" or "leg bracing" or "ankle brace*" or "ankle bracing" or "knee brace*" or "knee bracing" or "foot brace*" or "foot bracing" or "ankle tap*" or "knee tap*" or "leg tap*" or "foot tap*" or socks or heel NEXT/1 cup* or arch NEXT/1 support or "shoe insert*") AND (injury or injuries or sprain* or strain* or pain* or prevent* or prophyla*  or athletic or ankle or foot or "soft tissue*" or tendon* or leg or legs or hip or fasciitis or musculoskeletal or stress or fracture* or break*)) |
